# Heart disease mortality during the early pandemic period in the United States

**DOI:** 10.1101/2020.08.16.20175406

**Authors:** Jeremy Samuel Faust, Zhenqiu Lin, Kalen N. Wright, Michael A. Di Iorio, Carrie D. Walsh, Harlan M. Krumholz

## Abstract

**Importance:** The coronavirus disease 2019 (COVID-19) outbreak has been associated with decreases in acute myocardial infarction diagnoses (AMI) and admissions in the United States. Whether this affected heart disease deaths is unknown.

**Objective:** To determine whether changes in heart disease deaths occurred during the early pandemic period in the US, we analyzed areas without large COVID-19 outbreaks. This isolated the effect of decreased healthcare-seeking behavior during the early outbreak.

**Design, Setting, and Participants:** We performed an observational study of heart disease-specific mortality using National Center for Health Statistics data (NCHS). Weekly provisional counts were disaggregated by jurisdiction of occurrence during 2019 and 2020 for all-cause deaths, COVID-19 deaths, and heart disease deaths. For the primary analysis, jurisdictions were included if; 1) There was no all-cause excess mortality during the early pandemic period (weeks 14–17, 2020); 2) The completeness of that data was estimated by NCHS to be >97% as of July 22, 2020, and; 3) Decreases in emergency department (ED) visits occurred during the study period. We compared heart disease death rates during the early pandemic period with corresponding weeks in 2019 and a pre-pandemic control period of 2020 as a sensitivity analysis. Incident rate and rate ratios were calculated.

**Exposure:** The US COVID-19 outbreak.

**Main Outcomes and Measures:** Incidence of heart disease deaths.

**Results:** Twelve states met the primary inclusion criteria, capturing 747,375,188 person-weeks for the early pandemic period and 740,987,984 person-weeks for the 2019 control period. The mean incidence rate (per 100,000 person-weeks) for heart disease in states without excess deaths during the early pandemic period was 3.95 (95% CI 3.83 to 4.06) versus 4.19 (95% CI 4.14 to 4.23) during the corresponding period in 2019. The incident rate ratio (2020/2019) was 0.91 (95% CI 0.87 to 0.97). No state recorded an increase from either the corresponding period in 2019 or the 2020 prepandemic control period. Two states recorded fewer heart disease deaths.

**Conclusions and Relevance:** This observational study found a decrease in heart disease deaths during the early US outbreak in regions without significant COVID-19 burdens, despite decreases in ED utilization. Long term follow-up data are needed.

## Introduction

In the early phase of the COVID-19 pandemic, locations around the world reported a marked decrease in admissions for acute myocardial infarction (AMI), a phenomenon documented in the United States.^1–4^ In the United States, decreases in emergency department visits during this period were reported nationwide, regardless of whether a regional COVID-19 outbreak occurred.^5^–^6^

The principal concern is that the decline in AMI may have resulted from the avoidance of needed care.^7^ However, an alternative explanation is that changes in behaviors and the environment may have modified triggers for these events and decreased their incidence. If the decline was entirely avoidance of needed care, then we would expect to see an increase in cardiovascular mortality even in the short term. If there were an actual decline in incidence of cardiac emergencies, then we would expect to see a decline in cardiovascular mortality.

We, therefore, sought to determine the association between the decline in AMI admissions with cardiovascular death in areas with shelter-in-place orders but without many COVID-19 cases or overall excess deaths. Such an approach would reduce any misclassification of COVID-19 deaths. As a secondary analysis, we evaluated the association where there were excess deaths and many COVID-19 cases. We primarily analyzed weeks 14–17, 2020 because this corresponded to the largest decrease in emergency department use in a recent report.

## Methods

We performed an observational cohort study of heart disease-specific mortality by assembling publicly-available data from the United States Centers for Disease Control and Prevention’s National Center for Health Statistics (NCHS)^9^. Weekly provisional counts of all-cause deaths (by week that the deaths occurred) were reported by the NCHS, disaggregated by jurisdiction of occurrence, and for select underlying causes of death during 2019 and 2020, including COVID-19 deaths (ICD code UO71, Underlying Cause of Death), and heart disease deaths (ICD codes 100-109, 111, 113, 120-151).

For the primary analysis, we included U.S. jurisdictions that met three inclusion criteria; 1) There was no significant excess all-cause mortality during the early pandemic period (weeks 14–17) as adjudicated by the CDC; 2) The completeness of the mortality data was estimated by NCHS as being > 97% complete for weeks 14–17 as of July 22, 2020, and; 3) Decreases in emergency department visits occurred during the period of interest (see Supplemental Appendix Table 1).^10^ We compared heart disease death incidence rates during the early pandemic period (weeks 14–17) with the same weeks in 2019 and also to a pre-pandemic control period of 2020 (weeks 6–9) as a sensitivity analysis. States and other jurisdictions without excess mortality with estimated mortality data completeness of < 97% were deemed inadequately reliable for inclusion in the study.

We used SAS, version 9.4 (Cary, N.C.) for all analyses. Incidence rate per person-weeks and corresponding 95% confidence interval were calculated for weeks 14 through 17 for 2020 (early pandemic period) and the same weeks in 2019 (2019 control period) and weeks 6 through 9 2020 (pre-pandemic control period). To compare the heart-specific death incidence rate between the early pandemic period, the 2019 control, and the 2020 pre-pandemic control periods, we further calculated incidence rate ratios and their corresponding 95% confidence interval. The primary analysis captured 747,375,188 person-weeks for the early pandemic period and 101,620,248 person-weeks for the 2019 control period. As a secondary analysis, similar calculations were made for jurisdictions deemed by the CDC to have had statistically significant excess deaths during the early pandemic period, comprising 1,518,033,256 person-weeks in 2020 and 752,641,240 person-weeks in 2019. The sensitivity analysis compared the findings in the primary and secondary analysis to an additional 747,375,188 person-weeks of data during the 2020 pre-pandemic control period.

## Results

For the primary analysis, twelve states met the inclusion criteria. For the secondary analysis, 17 states and Washington, DC reported excess mortality in each of the four weeks of the early pandemic period. The mean incidence rate (per 100,000 person-weeks) for heart disease in states without excess deaths during the early pandemic period was 3.95 (95% CI 3.83 to 4.06) versus 4.35 (95% CI 4.23 to 4.48). The incident rate ratio was 0.91 (95% CI 0.87 to 0.97). In the primary analysis, heart disease deaths were unchanged in ten states and lower in two states when compared with the 2019 control period (incident rate ratios, Table 1A).

**Table 1A.**
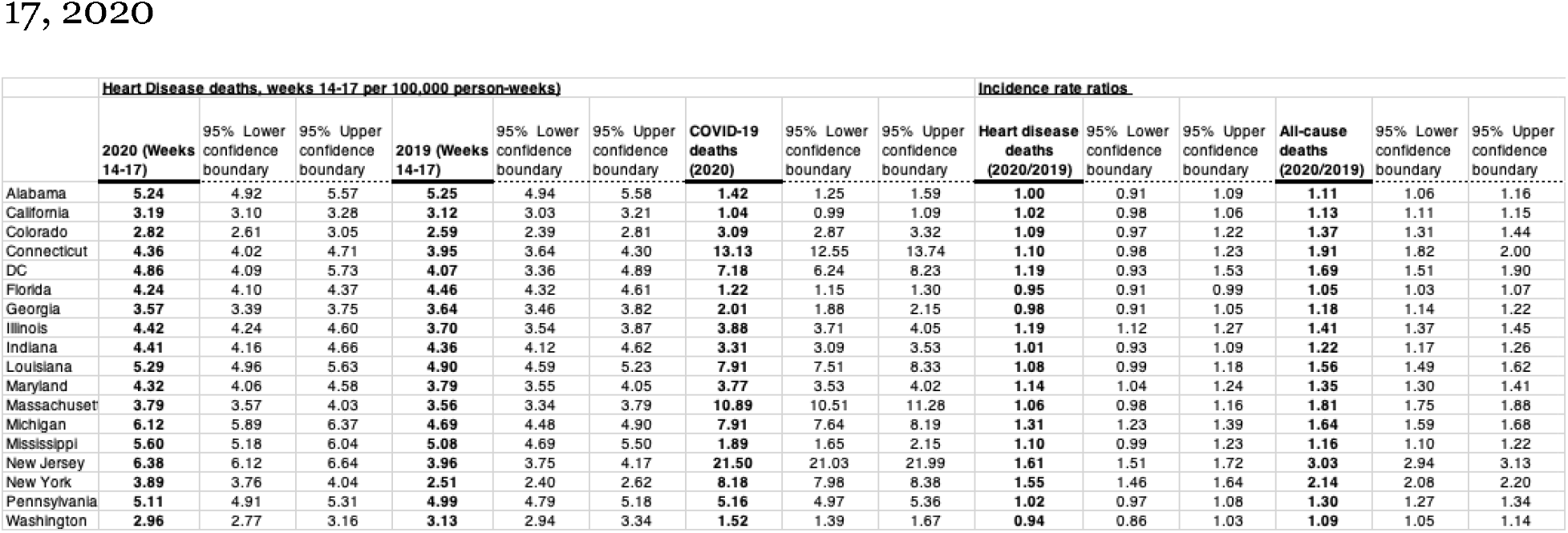
Primary Analysis. United States jurisdictions with no excess deaths with >97% estimated data completeness, weeks 14–17, 2020.

In the primary sensitivity analysis, heart disease death incident rates decreased during the pandemic versus pre-pandemic period (incident rate ratio 0.91; 95% CI 0.87 to 0.95) and there were no increases in any jurisdiction during the early pandemic period compared with the pre-pandemic control period (weeks 6–9, 2020); three individual states recorded statistically fewer heart disease deaths this year (Supplemental Appendix, Table 2A).

Overall in states with excess all-cause mortality (secondary analysis), the incident rate ratio for heart disease deaths was 1.12 (95% CI 1.10 to 1.13). Among the 18 jurisdictions in the secondary analysis, the incidence rate ratios of heart disease deaths were statistically higher in 5 of 18 jurisdictions as compared with the 2019 corresponding control period, lower in 1 jurisdiction, and unchanged in 12. (Table 1B). In the secondary sensitivity analysis, heart disease deaths were higher in 6 of 18 jurisdictions during the pandemic period as compared with the pre-pandemic control period, lower in 3 jurisdictions and unchanged in 9 (Supplemental appendix 2B) and overall the incident rate ratio was 1.06 (95% CI 1.05 to 1.08).

**Table 1B.**
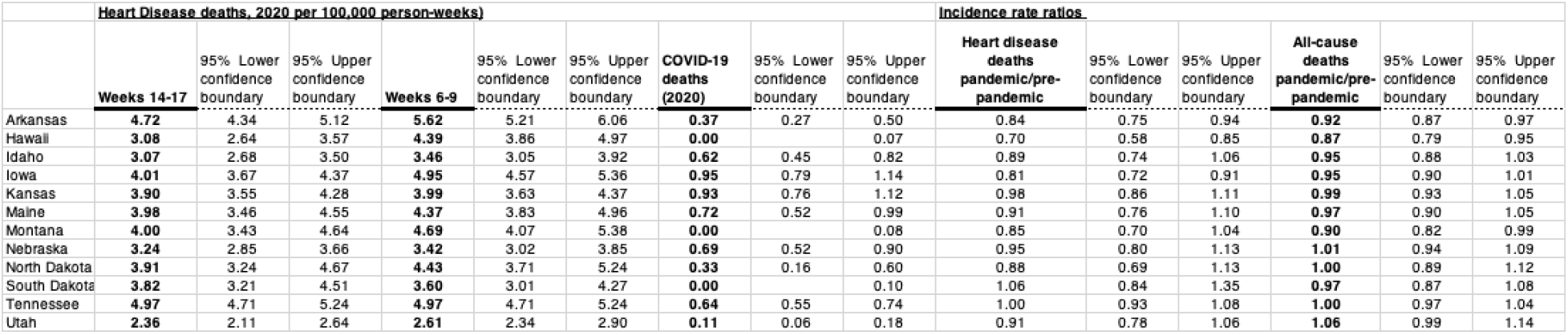
Secondary Analysis. United States jurisdictions with excess deaths, weeks 14-17, 2020

We observed an apparent correlation between an increase of heart disease deaths and all-cause death during the early pandemic period as compared with the 2019 control (Figure 1A) and the 2020 pre-pandemic control period (Figure 1B).

**Figure 1A (primary and secondary analyses) and 1B (sensitivity analyses).**
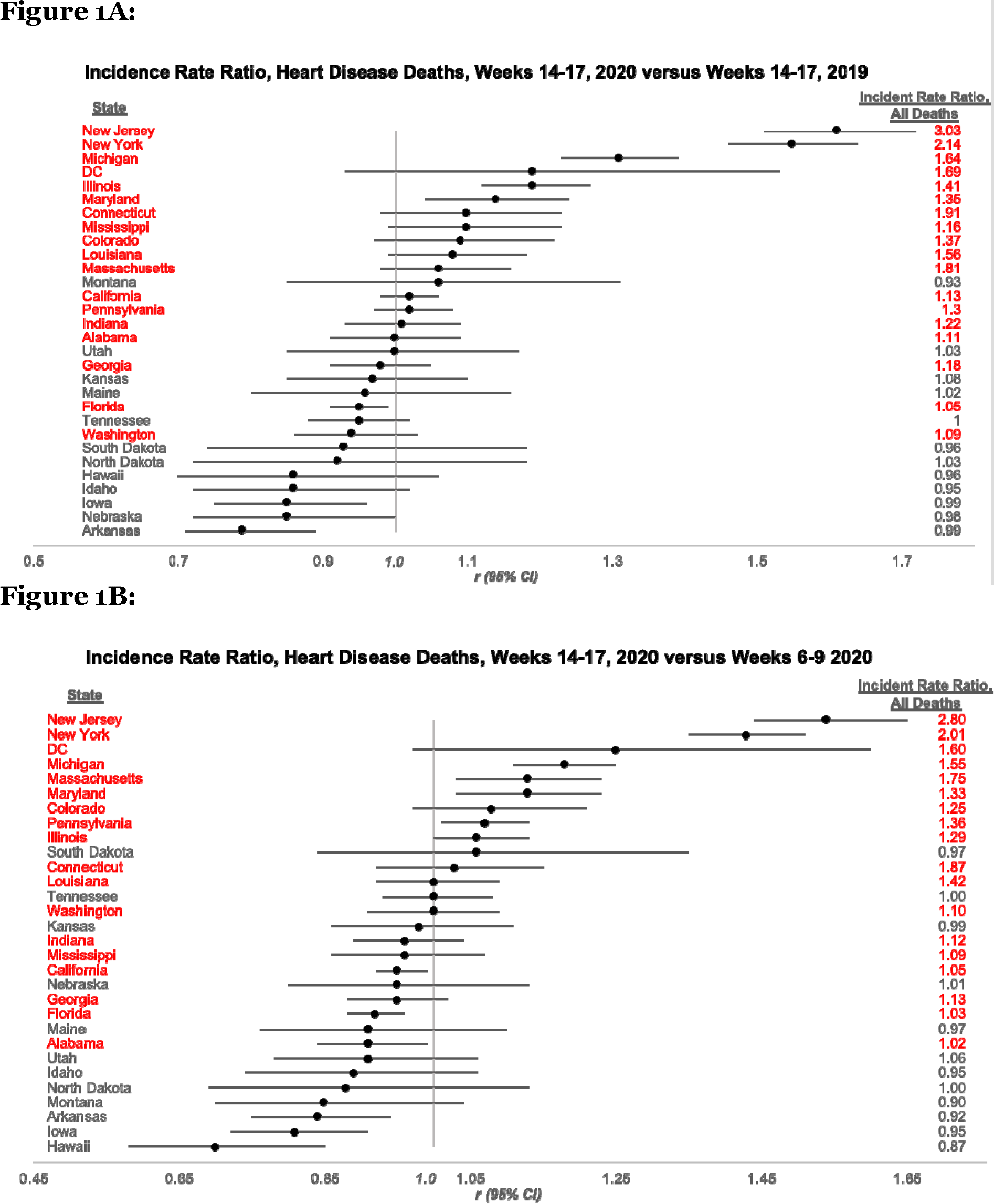
Incident rate ratios (IRR) point estimates for heart disease deaths are shown with their corresponding 95% confidence intervals. The vertical line indicates the boundary between increases in heart disease deaths (IRR > 1.0) and decreases (IRR < 1.0). Jurisdictions are organized with areas of largest increase in heart disease deaths in the top row, descending to areas with the largest measured decreases at the bottom. IRRs for all-cause mortality are shown at the right to provide context for COVID-19 outbreak during the study period. Red indicates states deemed by the CDC to have reached the threshold for excess mortality during the entire study period.

## Discussion

This study provides evidence that the reported decrease in AMI admissions was not associated with a discernible short-term increase in cardiovascular deaths, but rather with a decline. We were able to isolate the effect of the change in healthcare utilization because our primary analysis focused on areas of the country that experienced decreases in emergency department use but had not yet experienced a substantial enough COVID-19 outbreak to have caused an increase in all-cause mortality. In contrast, there was an increase in cardiovascular deaths in some areas where there were increases in mortality due to COVID-19, which may represent a misclassification of deaths in these regions or an association between COVID-19 itself and the risk of cardiovascular deaths.

There are limitations to this study. While death certificates have limitations, we compared death certificate information in different time periods but within the same jurisdictions. Therefore, any biases should be generally consistent. The design cannot determine what caused the change in heart disease-specific mortality in the regions which experienced significant COVID-19 burdens. We also cannot determine long-term outcomes at this stage, and some of the harm of avoidance in care may only be manifest over time.

The study provides important new information suggesting that changes in behaviors and exposures in non-COVID-19 areas may have, in part, reduced triggers for acute cardiovascular events. There may have also been some harm with avoidance of care. In some areas with large COVID-19 burdens, cardiovascular deaths increased; future research needs to untangle whether there is misclassification during a period of excess deaths or whether COVID-19 itself increases cardiovascular deaths.

## Data Availability

All data are available to the public

## Funding Statement

### Data Transparency Statement

Drs. Faust, Lin, Wright and Krumholz had access to the entire dataset at all times. All data are available to the public via the National Center for Health Statistics.

Drs. Walsh and Di Iorio contributed data used for statistical analysis and inclusion criteria determination.

## Question

Did heart disease death rates change during the early pandemic period in the United States?

## Findings

In this observational cohort study of public data from the National Center for Health Statistics, we found that in U.S. states without excess mortality during the early COVID-19 outbreak, heart disease deaths decreased during the early pandemic period (weeks 14–17, 2020), despite previously described widespread decreases in acute myocardial infarctions and emergency department visits during this time. No states reported an increase in heart disease specific death as compared to the corresponding period in 2019 and a 2020 pre-pandemic control period.

## Meaning

In areas without large COVID-19 outbreaks, previously described decreases in healthcare seeking behavior and acute myocardial infarction diagnoses during the early pandemic period were associated with a decrease in heart disease-specific mortality in the short term.

## Supplemental Appendix

1. Supplemental Appendix Table 1. (p.1)
2. Supplemental Appendix Table 2A and 2B (p.2)

**Table S1.**
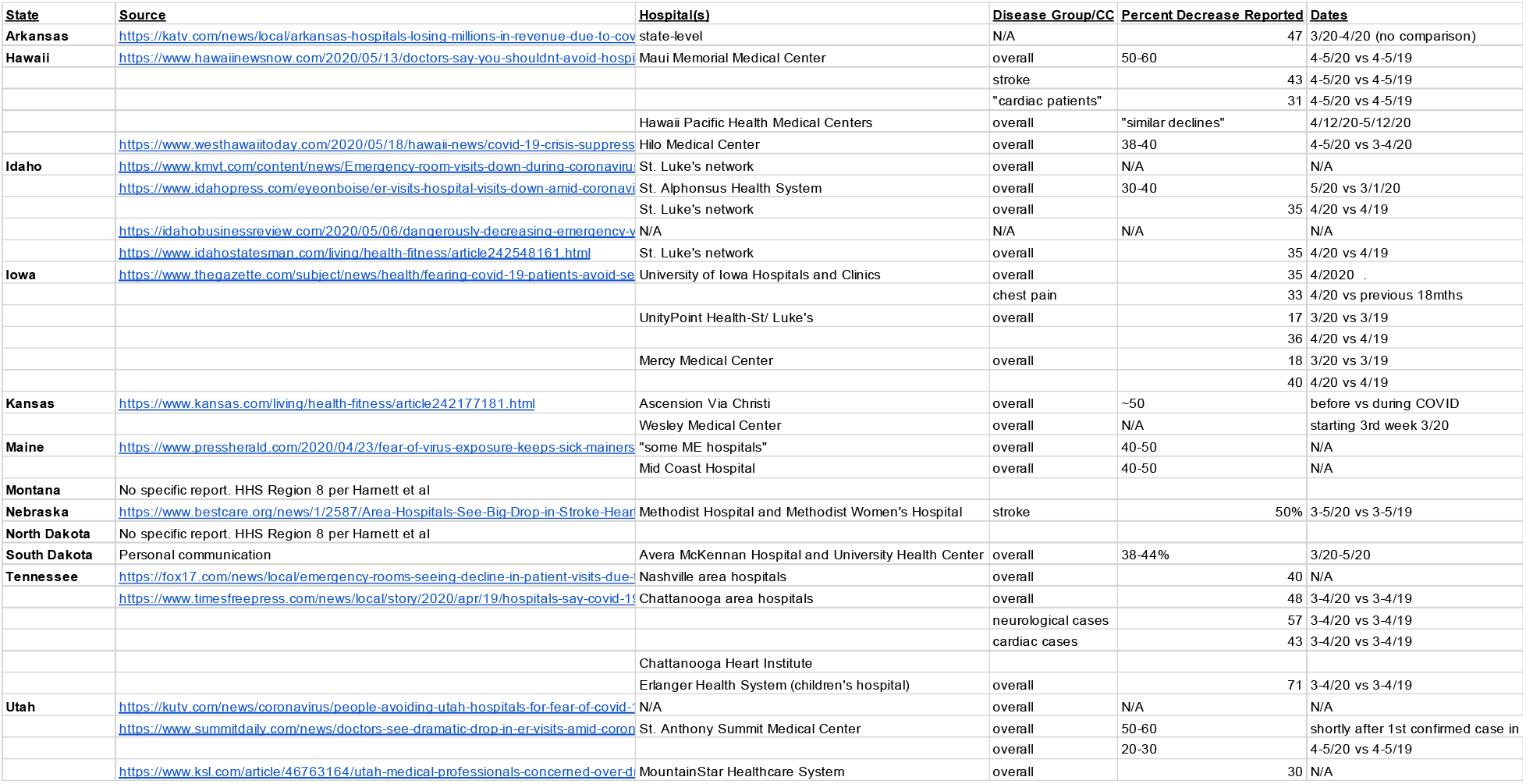
Reports of decreases in emergency department visits among states included in the primary analysis. Montana and North Dakota were included in the primary analysis because it was determined based on CDC disaggregated data (HHS Region 8) that these regions were likely to have had some decrease in emergency department visits. Exclusion of these states would not change the qualitative findings we report.

**Table S2A.**
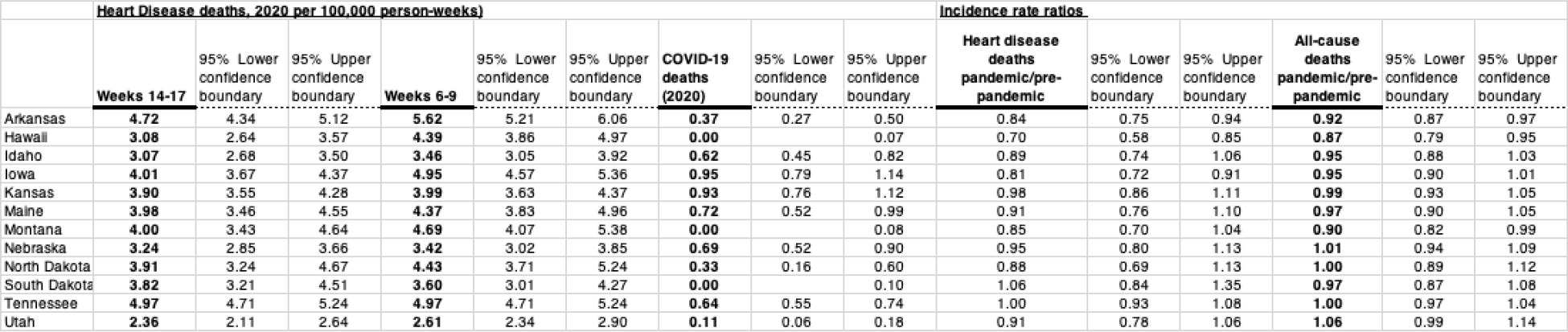
Sensitivity analysis. United States jurisdictions with no excess deaths with >97% estimated data completeness, weeks 6–9, 2020.

**Table S2B.** Sensitivity analysis. United States jurisdictions with excess deaths, weeks 6–9, 2020.

|  | Heart Disease deaths, 2020 per 100,000 person-weeks) |  |  |  |  |  | Incidence rate ratios |  |  |  |  |  |  |  |  |
| --- | --- | --- | --- | --- | --- | --- | --- | --- | --- | --- | --- | --- | --- | --- | --- |
|  |  | 95% Lower confidence boundary | 95% Upper confidence boundary |  | 95% Lower confidence boundary | 95% Upper confidence boundary | COVID-19 deaths (2020) | 95% Lower confidence boundary | 95% Upper confidence boundary | Heart disease deaths pandemic/pre-pandemic | 95% Lower confidence boundary | 95% Upper confidence boundary | All-cause deaths pandemic/pre-pandemic | 95% Lower confidence boundary | 95% Upper confidence boundary |
|  | Weeks 14-17 |  |  | Weeks 6-9 |  |  |  |  |  |  |  |  |  |  |  |
| Alabama | 5.24 | 4.92 | 5.57 | 5.76 | 5.42 | 6.10 | 1.42 | 1.25 | 1.59 | 0.91 | 0.84 | 0.99 | 1.02 | 0.97 | 1.06 |
| California | 3.19 | 3.10 | 3.28 | 3.34 | 3.25 | 3.43 | 1.04 | 0.99 | 1.09 | 0.95 | 0.92 | 0.99 | 1.05 | 1.03 | 1.07 |
| Colorado | 2.82 | 2.61 | 3.05 | 2.61 | 2.41 | 2.82 | 3.09 | 2.87 | 3.32 | 1.08 | 0.97 | 1.21 | 1.25 | 1.20 | 1.31 |
| Connecticut | 4.36 | 4.02 | 4.71 | 4.24 | 3.91 | 4.60 | 13.13 | 12.55 | 13.74 | 1.03 | 0.92 | 1.15 | 1.87 | 1.78 | 1.96 |
| DC | 4.86 | 4.09 | 5.73 | 3.89 | 3.20 | 4.67 | 7.18 | 6.24 | 8.23 | 1.25 | 0.97 | 1.60 | 1.60 | 1.43 | 1.79 |
| Florida | 4.24 | 4.10 | 4.37 | 4.62 | 4.48 | 4.77 | 1.22 | 1.15 | 1.30 | 0.92 | 0.88 | 0.96 | 1.03 | 1.00 | 1.05 |
| Georgia | 3.57 | 3.39 | 3.75 | 3.77 | 3.59 | 3.96 | 2.01 | 1.88 | 2.15 | 0.95 | 0.88 | 1.02 | 1.13 | 1.09 | 1.16 |
| Illinois | 4.42 | 4.24 | 4.60 | 4.15 | 3.98 | 4.34 | 3.88 | 3.71 | 4.05 | 1.06 | 1.00 | 1.13 | 1.29 | 1.25 | 1.32 |
| Indiana | 4.41 | 4.16 | 4.66 | 4.59 | 4.34 | 4.85 | 3.31 | 3.09 | 3.53 | 0.96 | 0.89 | 1.04 | 1.12 | 1.08 | 1.16 |
| Louisiana | 5.29 | 4.96 | 5.63 | 5.29 | 4.96 | 5.63 | 7.91 | 7.51 | 8.33 | 1.00 | 0.92 | 1.09 | 1.42 | 1.37 | 1.48 |
| Maryland | 4.32 | 4.06 | 4.58 | 3.83 | 3.58 | 4.08 | 3.77 | 3.53 | 4.02 | 1.13 | 1.03 | 1.23 | 1.33 | 1.28 | 1.39 |
| Massachusetts | 3.79 | 3.57 | 4.03 | 3.36 | 3.15 | 3.58 | 10.89 | 10.51 | 11.28 | 1.13 | 1.03 | 1.23 | 1.75 | 1.69 | 1.81 |
| Michigan | 6.12 | 5.89 | 6.37 | 5.19 | 4.97 | 5.41 | 7.91 | 7.64 | 8.19 | 1.18 | 1.11 | 1.25 | 1.55 | 1.50 | 1.59 |
| Mississippi | 5.60 | 5.18 | 6.04 | 5.84 | 5.41 | 6.29 | 1.89 | 1.65 | 2.15 | 0.96 | 0.86 | 1.07 | 1.09 | 1.03 | 1.15 |
| New Jersey | 6.38 | 6.12 | 6.64 | 4.13 | 3.93 | 4.35 | 21.50 | 21.03 | 21.99 | 1.54 | 1.44 | 1.65 | 2.80 | 2.72 | 2.88 |
| New York | 3.89 | 3.76 | 4.04 | 2.72 | 2.61 | 2.84 | 8.18 | 7.98 | 8.38 | 1.43 | 1.35 | 1.51 | 2.01 | 1.96 | 2.07 |
| Pennsylvania | 5.11 | 4.91 | 5.31 | 4.78 | 4.59 | 4.97 | 5.16 | 4.97 | 5.36 | 1.07 | 1.01 | 1.13 | 1.36 | 1.32 | 1.39 |
| Washington | 2.96 | 2.77 | 3.16 | 2.96 | 2.77 | 3.16 | 1.52 | 1.39 | 1.67 | 1.00 | 0.91 | 1.09 | 1.10 | 1.06 | 1.14 |

